# Missed Opportunities for Stroke Prevention in Hypertensive Patients: A Retrospective Case-Control Study

**DOI:** 10.64898/2026.04.21.26351407

**Authors:** Huanhuan Yang, Yuntian Liu, Chungsoo Kim, Chenxi Huang, Mitsuaki Sawano, Patrick Young, Mark Anderson, John S. Burrows, Harlan M. Krumholz, John E. Brush, Yuan Lu

**Affiliations:** Center for Outcomes Research and Evaluation, Yale New Haven Hospital, New Haven, CT, USA; Vanke School of Public Health, Tsinghua University, Beijing, China; Section of Cardiovascular Medicine, Department of Medicine, Yale School of Medicine, New Haven, CT, USA; Department of Biomedical Informatics & Data Science, Yale School of Medicine, New Haven, CT, USA; Department of Biomedical Informatics, Ajou University School of Medicine, Suwon, Republic of Korea; Teikyo Academic Research Center, Teikyo University, Tokyo, Japan; Sentara Health, Norfolk, VA, USA; Department of Health Policy and Management, Yale School of Public Health, New Haven, Connecticut; Macon & Joan Brock Virginia Health Sciences at Old Dominion University, Norfolk, VA, USA

**Author notes:** **Corresponding authors:** John E. Brush, Jr., MD, 800 Independence Blvd, Virginia Beach, VA, 23455. & Yuan Lu, ScD, 195 Church Street, Fifth Floor, New Haven, CT, 06510. Contributed equally as Co-First Authors (Huanhuan Yang, Yuntian Liu).

**Keywords:** Blood Pressure, Case-Control Study, Hypertension, Ischemic Stroke, Missed Opportunities, Treatment Intensification

## Abstract

**Background:** Hypertension is the leading modifiable risk factor for ischemic stroke, yet the adequacy of preventative hypertension care in routine clinical practice remains suboptimal. Whether gaps in hypertension management represent missed opportunities for stroke prevention remains unclear.

**Objective:** To evaluate the association between hypertension care delivery and the risk of incident ischemic stroke.

**Methods:** We conducted a retrospective, matched, nested case-control study among adults with hypertension using electronic health record data from a large regional health system (2010–2024). Patients with a first-ever ischemic stroke were matched 1:2 to controls on age, sex, race and ethnicity, and calendar time. Three care metrics were assessed during follow-up: (1) outpatient visits with blood pressure (BP) measurement per year; (2) number of antihypertensive medication ingredients; and (3) medication intensification score. Conditional logistic regression estimated adjusted odds ratios (aORs).

**Results:** The study included 13,476 cases and 26,952 matched controls (N = 40,428). Mean (SD) age was 64.8 (12.2) years, 54.1% were female, and mean follow-up was 2,497 (1,308) days. Cases had fewer BP visits per year (median, 2.50 vs. 3.01; p < 0.001), similar number of medication ingredients (2.00 vs 2.00), and lower treatment intensification scores (-0.211 vs -0.125). In adjusted models, >5 BP visits per year was associated with lower stroke odds (aOR, 0.55; 95% CI, 0.51-0.59) compared with ≤1 visit. Use of 2-3 medication ingredients (vs 0) was also associated with reduced stroke odds (aOR, 0.80; 95% CI, 0.75-0.86), whereas >3 ingredients was not significant. The highest quartile of treatment intensification showed the strongest association (aOR, 0.47; 95% CI, 0.44-0.51). Findings were consistent across subgroup and sensitivity analyses, including strata defined by baseline SBP and follow-up SBP.

**Conclusions:** Greater engagement in hypertension care was associated with lower odds of ischemic stroke, suggesting that gaps in routine management may represent missed opportunities for prevention.

**Novelty and Relevance:** *What Is New?:*

- In this nested case-control study, differences in hypertension care over time—including follow-up frequency, medication use, and treatment adjustments—were associated with incident ischemic stroke.

*What Is Relevant?:*

- Variations in hypertension care delivery over time may represent missed opportunities for stroke prevention regardless of baseline or average SBP levels during follow-up.
- Improving follow-up and timely treatment adjustments may offer actionable targets for primary prevention.

*Clinical/Pathophysiological Implications:*

- Longitudinal gaps in hypertension management may contribute to stroke risk beyond BP control, highlighting opportunities to improve prevention through more consistent care delivery.

## Introduction

Each year in the United States, approximately 795,000 individuals experience a new or recurrent stroke, of which nearly 87% are ischemic strokes.^1^ Stroke is the third major cause of death and long-term disability worldwide.^2^ Importantly, elevated systolic blood pressure (SBP) accounts for more than one-half of the population-attributable risk for stroke.^2^ Thus, lowering SBP is an important and highly effective strategy for reducing stroke risk.^3,4^

However, hypertension management in routine clinical practice remains suboptimal. National data indicate that fewer than half of US adults with hypertension achieve SBP <140/90 mm Hg, and fewer than one-quarter meet more stringent <130/80 mm Hg targets.^3,4^ Achieving and maintaining BP control in practice requires sustained engagement with clinical care, including regular follow-up visits, appropriate medication use, and timely treatment intensification. These elements collectively reflect the delivery of hypertension care over time. However, it remains unclear whether differences in hypertension care delivery translate into meaningful differences in clinical outcomes.

Prior studies have described “therapeutic inertia”, defined as the failure to initiate or intensify antihypertensive therapy when BP is uncontrolled, and have documented its high prevalence across diverse clinical settings.^5,6^ However, most investigations of therapeutic inertia have focused on intermediate outcomes such as BP control rather than unambiguous cardiovascular endpoints such as ischemic stroke.^7^ Furthermore, approaches to quantifying treatment intensification vary substantially across studies and may incompletely capture real-world therapeutic responsiveness to elevated BP.^8^ More broadly, few studies have systematically characterized longitudinal hypertension care patterns, including follow-up frequency, antihypertensive medication exposure, and treatment intensification, as a unified care delivery construct in relation to stroke risk in routine practice. In addition, differences in observation windows or follow-up duration between comparison groups in observational studies may complicate interpretation of the temporal relationship between management gaps and subsequent stroke events.

To address these gaps, we leveraged longitudinal electronic health record (EHR) data standardized to the Observational Medical Outcomes Partnership (OMOP) common data model to examine patterns of hypertension care preceding stroke. Using a time-aligned matched case-control design to ensure comparable observation windows, we quantified the association between hypertension care delivery and the risk of incident ischemic stroke. By linking real-world patterns of BP management to stroke outcomes, this study aims to identify potentially modifiable gaps in hypertension care that may represent missed opportunities for stroke prevention.

## Methods

### Data Source

This retrospective case-control study was conducted using EHR data from Sentara Health, a large nonprofit integrated regional healthcare system serving Virginia and northeastern North Carolina, comprising 12 hospitals, 500+ outpatient care sites, and affiliated medical groups. All EHR data were standardized to the OMOP common data model. The study period spanned from January 1, 2010, to December 31, 2024.

### Study Population

The study population was derived from 3,728,562 patients with at least one documented encounter during the study period (Supplemental Figure 1). To ensure adequate longitudinal observation, we restricted the cohort to patients with documented encounters over at least 5 years, defined as the interval between the first and last recorded encounter (n = 1,390,427 patients). To ensure adequate frequency of observations, we then restricted the cohort to patients with a rate of years with visits (RYV) ≥0.5. RYV was calculated as the number of calendar years with at least one documented encounter divided by the total calendar years of the observation period. Among the remaining patients (n = 1,027,277), we identified 546,120 individuals with hypertension, forming the hypertensive cohort. The definition of hypertension is provided below.

### Study Design

We conducted a retrospective 1:2 matched nested case-control study within the hypertensive cohort (Figure 1). The index date (T□ for cases and T□′ for controls) was defined as the date when patients first met the definition of hypertension. Cases were defined as patients with an incident ischemic stroke occurring more than 60 days after T□. A 60-day blanking period was applied to reduce potential reverse causation and to allow sufficient time for exposure assessment. Care processes occurring within 60 days after the index date were excluded, as healthcare encounters shortly after hypertension diagnosis may reflect diagnostic evaluation or early clinical management rather than stable patterns of routine hypertension care.^9^ The stroke admission date was defined as T□. For each case, the duration between T□ and T□ (ΔT) was calculated. Controls were selected from patients in the hypertensive cohort who remained free of stroke during the corresponding follow-up interval.

**Figure 1.**
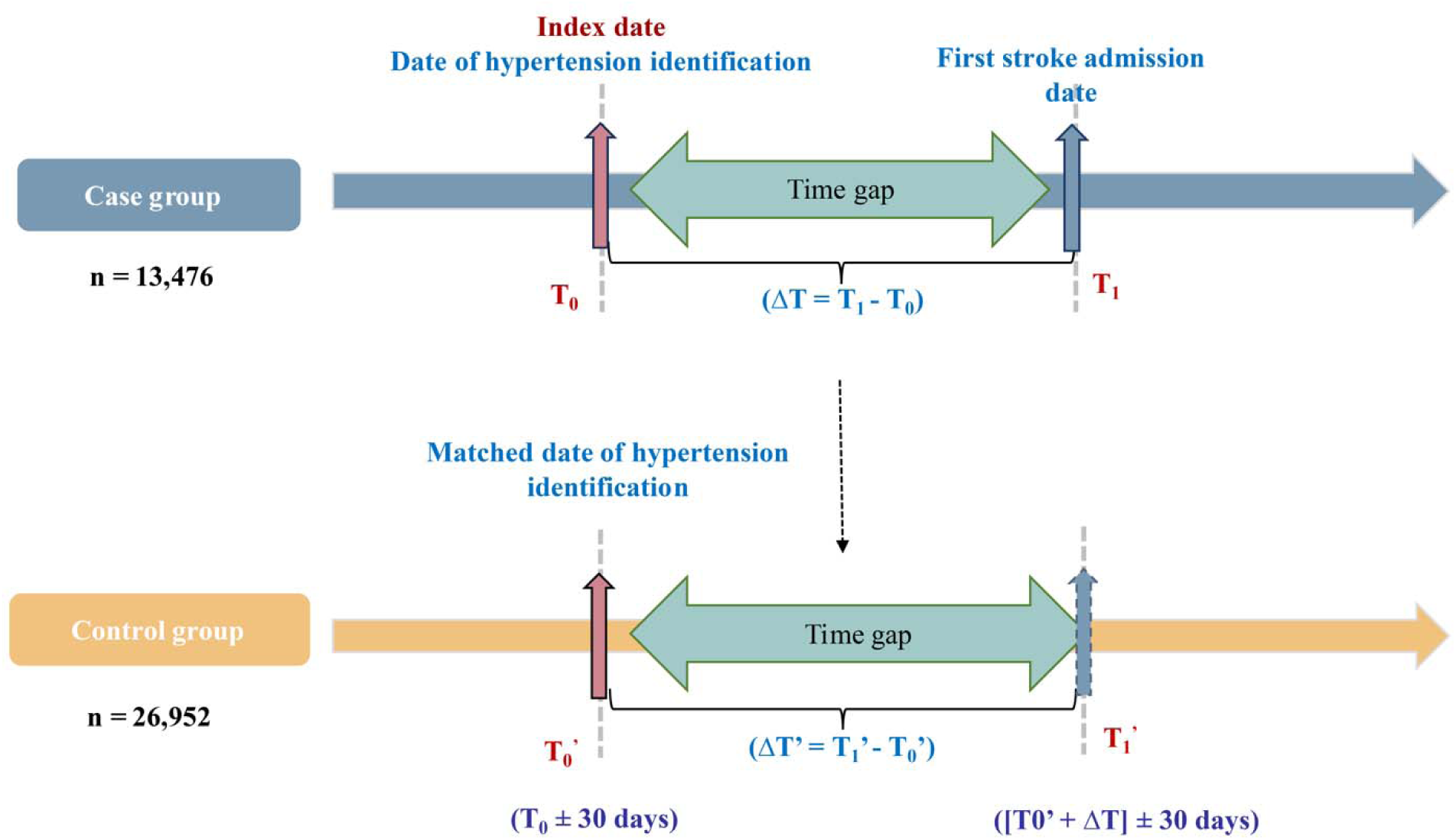
Study design and time-aligned matching strategy.

### Matching Strategy and Temporal Alignment

Each case was matched with two controls (1:2 matching) on age, sex, and race/ethnicity at T□ the index date. To ensure temporal comparability, controls were additionally required to have index date (T□′) within ±30 days of the corresponding case’s T□. Among eligible controls, those with the smallest absolute difference in index date were preferentially selected. Each control patient was selected only once. To align follow-up duration between cases and controls, we implemented a time-aligned matching approach. For each matched control, a pseudo-event date (T□′) was generated by adding the case-specific ΔT to the control patient’s index date (T□′). This ensured identical observation windows (T□ to T□ for cases; T□′ to T□′ for controls). To minimize bias due to differential follow-up or informative loss to care, controls were required to have at least one clinical encounter within ±30 days of T□, indicating ongoing engagement in care at the comparable time point.After matching and temporal alignment, matched sets without outpatient visits with recorded BP measurements during the observation window were excluded. The final analytic cohort included a total of 40,428 participants (13,476 cases and 26,952 controls).

### Definition of Hypertension

Hypertension was defined by the presence of any one of the following: (1) a coded diagnosis of hypertension (Supplemental Table 1); (2) a recorded prescription for one or more antihypertensive medications (Supplemental Table 2); or (3) at least two outpatient visits with elevated BP (either SBP ≥140 or diastolic BP [DBP] ≥90 mmHg). The 140/90 mmHg threshold was used to identify clearly elevated BP consistent with clinical practice and hypertension definitions widely used during most of the study period (2010–2024). If multiple BP measurements were recorded during a visit, the first reading was discarded, and the mean of the remaining measurements was calculated.^10^ If multiple outpatient visits occurred on the same day, the average BP across visits was calculated.

### Definition of Ischemic Stroke

Ischemic stroke was defined as an incident ischemic cerebrovascular event identified using validated OMOP concept sets (Supplemental Table 1). Patients with hemorrhagic stroke were excluded. If ischemic stroke and hemorrhagic stroke were both recorded on the same day, the event was classified as an ischemic stroke.

### Definition of Hypertension Care Delivery

Hypertension care delivery during the observation window (T□ to T□ for cases and T□′ to T□′ for controls) was quantified using three complementary measures: number of outpatient visits with BP measurement per year; number of antihypertensive medication ingredients; antihypertensive medication intensification score.

The antihypertensive medication intensification score was calculated as:^11^

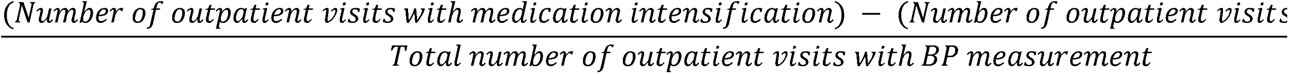

The score ranges from -1.0 to 1.0. Higher values reflect greater therapeutic responsiveness relative to episodes of uncontrolled BP, whereas lower values indicate a higher burden of elevated BP relative to medication intensification. Elevated BP used for defining treatment intensification was based on a threshold of SBP ≥140 mmHg or DBP ≥90 mmHg. This threshold was selected to identify episodes of clearly uncontrolled hypertension where treatment escalation would typically be expected in routine clinical practice.

Medication intensification was defined as 1) initiation of antihypertensive therapy (transition from untreated to treated), 2) addition of a new antihypertensive medication ingredient, or 3) an increase in ingredient-level daily dose compared with the prior visit. All intensification events occurring within the observation window were counted, regardless of whether elevated BP was documented at the same visit.

### Covariates

Baseline covariates assessed at T□ included age, sex, race and ethnicity, area-level social vulnerability index (SVI) based on the patient’s recorded address at baseline, baseline SBP and DBP, and comorbid conditions identified prior to T□, including congestive heart failure (CHF), chronic kidney disease (CKD), chronic obstructive pulmonary disease (COPD), atrial fibrillation (AF), diabetes, and dementia (Supplemental Table 1)

### Statistical Analysis

Baseline characteristics were summarized using means (SD) or median (interquartile range, IQR) for continuous variables, and frequencies (%) for categorical variables. Group differences were assessed using Welch’s two-sample t-tests or Wilcoxon rank-sum tests for continuous variables and Pearson’s chi-square tests for categorical variables, as appropriate. Given the matched nested case-control design with temporally aligned observation windows, associations between hypertension care delivery measures and incident ischemic stroke were estimated using conditional logistic regression to account for the matched design.^12^ Three models were constructed: Model 0 was unadjusted; Model 1 was adjusted for baseline SBP; Model 2 was further adjusted for SVI, CHF, CKD, COPD, AF, diabetes, and dementia. Care delivery measures were modeled as categorical variables according to pre-specified groupings: outpatient visits with BP measurement per year (≤1, 2-3, 4-5, >5 visits), number of antihypertensive medication ingredients (0, 1, 2-3, >3), and treatment intensification score (quartiles). Categorization was chosen to facilitate clinical interpretability. Pre-specified subgroup analyses were conducted stratified by baseline SBP, average SBP during the observation window (<130 vs ≥130 mmHg), sex, age group, race/ethnicity, and SVI (quartile). Interaction terms were tested to evaluate effect modification. In sensitivity analyses, a more stringent threshold (BP ≥160/100 mmHg) was applied. Additional analyses were conducted after excluding matched sets in which any patient had baseline cardiometabolic comorbidities (CHF, AF, or CKD), and in a separate analysis after excluding matched sets with any baseline comorbidity (CHF, AF, or CKD, COPD, diabetes, or dementia). Missing data were handled through study design. Participants with missing key variables were excluded during cohort construction and matching.

All statistical tests were two-sided, with P <0.05 considered statistically significant. Data extraction from the OMOP common data model was performed using Microsoft SQL Server, and statistical analyses were conducted using R version 4.2.3 (R Foundation for Statistical Computing).

The study protocol was approved by the Institutional Review Board of the Malcolm and Joan Brock Virginia Health Sciences at Old Dominion University, which granted a waiver of informed consent and authorization due to the retrospective design. The study was conducted in accordance with STROBE guidelines.

## Results

Basic characteristics were shown in Table 1. Age, sex, race and ethnicity, and follow-up duration (mean [SD], 2,497.4 [1,307.6] days) were well balanced between cases and controls. Mean (SD) age at index was 64.8 (12.2) years, and 54.1% were women. The cohort was predominantly non-Hispanic White (64.9%) and non-Hispanic Black (33.8%). Distribution of the SVI differed between cases and controls (p < 0.001), with cases slightly more likely to reside in more vulnerable areas. Baseline SBP and DBP were higher among cases compared with controls (SBP, 135.7 vs. 132.1 mm Hg; DBP, 77.0 vs. 76.5 mm Hg; both p < 0.001). Average follow-up BP values were also higher in cases (SBP, 135.5 vs. 131.6 mm Hg; DBP, 76.2 vs. 75.4 mm Hg; both p < 0.001). Prevalence of several comorbidities, including CHF, CKD, COPD, AF, diabetes, and dementia, was also higher among cases (all p < 0.001).

**Table 1.**
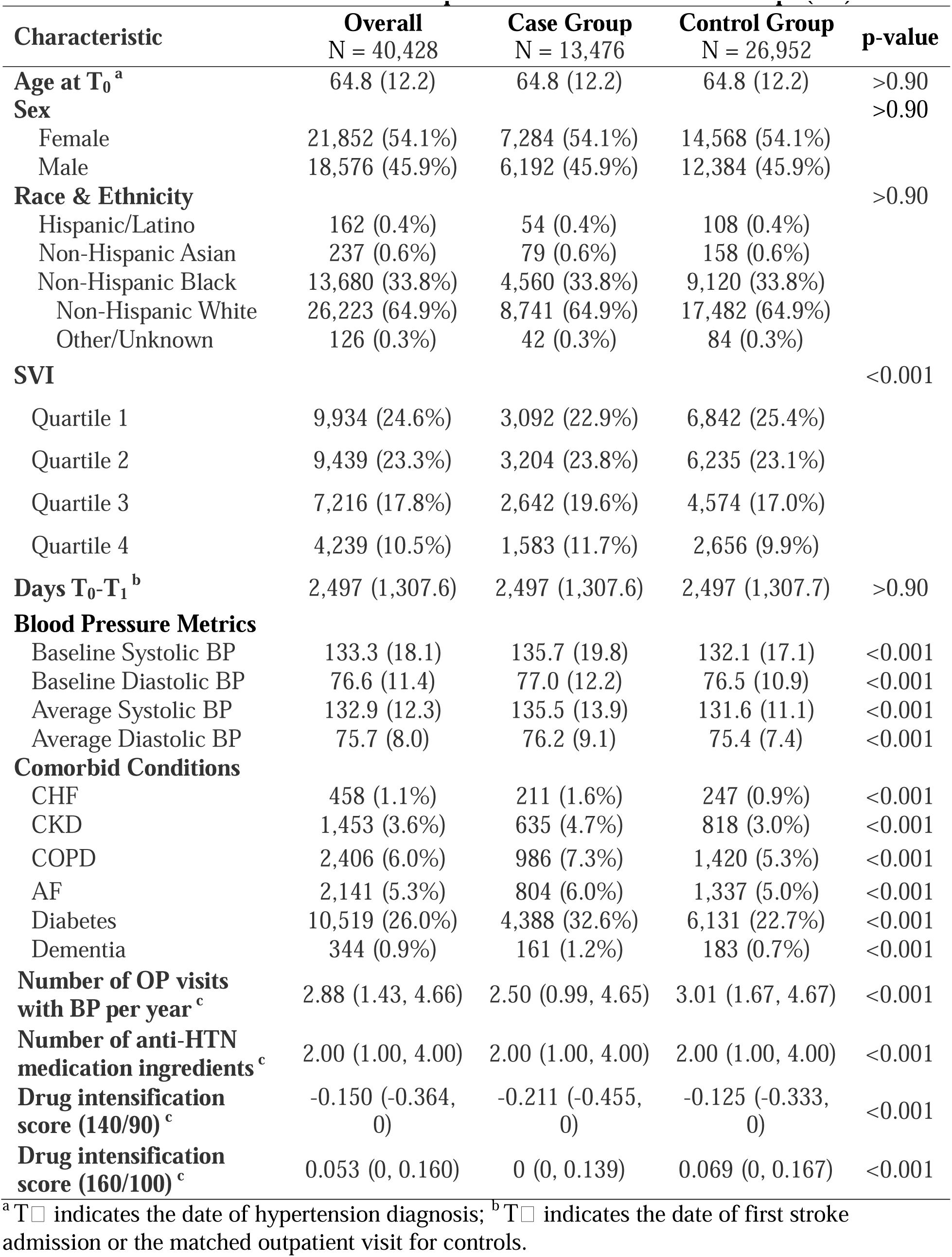
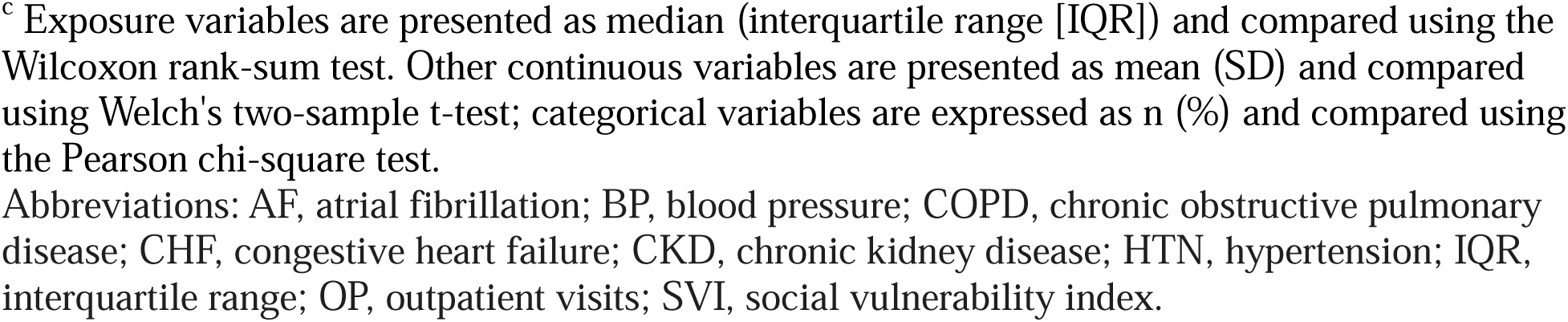
Baseline Characteristics of Participants in Case and Control Groups (1:2).

| Characteristic | Overall<br>N = 40,428 | Case Group<br>N = 13,476 | Control Group<br>N = 26,952 | p-value |
| --- | --- | --- | --- | --- |
| **Age at T<sub>0</sub><sup>a</sup>** | 64.8 (12.2) | 64.8 (12.2) | 64.8 (12.2) | >0.90 |
| **Sex** |  |  |  | >0.90 |
| Female | 21,852 (54.1%) | 7,284 (54.1%) | 14,568 (54.1%) |  |
| Male | 18,576 (45.9%) | 6,192 (45.9%) | 12,384 (45.9%) |  |
| **Race & Ethnicity** |  |  |  | >0.90 |
| Hispanic/Latino | 162 (0.4%) | 54 (0.4%) | 108 (0.4%) |  |
| Non-Hispanic Asian | 237 (0.6%) | 79 (0.6%) | 158 (0.6%) |  |
| Non-Hispanic Black | 13,680 (33.8%) | 4,560 (33.8%) | 9,120 (33.8%) |  |
| Non-Hispanic White | 26,223 (64.9%) | 8,741 (64.9%) | 17,482 (64.9%) |  |
| Other/Unknown | 126 (0.3%) | 42 (0.3%) | 84 (0.3%) |  |
| **SVI** |  |  |  | <0.001 |
| Quartile 1 | 9,934 (24.6%) | 3,092 (22.9%) | 6,842 (25.4%) |  |
| Quartile 2 | 9,439 (23.3%) | 3,204 (23.8%) | 6,235 (23.1%) |  |
| Quartile 3 | 7,216 (17.8%) | 2,642 (19.6%) | 4,574 (17.0%) |  |
| Quartile 4 | 4,239 (10.5%) | 1,583 (11.7%) | 2,656 (9.9%) |  |
| **Days T<sub>0</sub>-T<sub>1</sub><sup>b</sup>** | 2,497 (1,307.6) | 2,497 (1,307.6) | 2,497 (1,307.7) | >0.90 |
| **Blood Pressure Metrics** |  |  |  |  |
| Baseline Systolic BP | 133.3 (18.1) | 135.7 (19.8) | 132.1 (17.1) | <0.001 |
| Baseline Diastolic BP | 76.6 (11.4) | 77.0 (12.2) | 76.5 (10.9) | <0.001 |
| Average Systolic BP | 132.9 (12.3) | 135.5 (13.9) | 131.6 (11.1) | <0.001 |
| Average Diastolic BP | 75.7 (8.0) | 76.2 (9.1) | 75.4 (7.4) | <0.001 |
| **Comorbid Conditions** |  |  |  |  |
| CHF | 458 (1.1%) | 211 (1.6%) | 247 (0.9%) | <0.001 |
| CKD | 1,453 (3.6%) | 635 (4.7%) | 818 (3.0%) | <0.001 |
| COPD | 2,406 (6.0%) | 986 (7.3%) | 1,420 (5.3%) | <0.001 |
| AF | 2,141 (5.3%) | 804 (6.0%) | 1,337 (5.0%) | <0.001 |
| Diabetes | 10,519 (26.0%) | 4,388 (32.6%) | 6,131 (22.7%) | <0.001 |
| Dementia | 344 (0.9%) | 161 (1.2%) | 183 (0.7%) | <0.001 |
| **Number of OP visits with BP per year<sup>c</sup>** | 2.88 (1.43, 4.66) | 2.50 (0.99, 4.65) | 3.01 (1.67, 4.67) | <0.001 |
| **Number of anti-HTN medication ingredients<sup>c</sup>** | 2.00 (1.00, 4.00) | 2.00 (1.00, 4.00) | 2.00 (1.00, 4.00) | <0.001 |
| **Drug intensification score (140/90)<sup>c</sup>** | -0.150 (-0.364, 0) | -0.211 (-0.455, 0) | -0.125 (-0.333, 0) | <0.001 |
| **Drug intensification score (160/100)<sup>c</sup>** | 0.053 (0, 0.160) | 0 (0, 0.139) | 0.069 (0, 0.167) | <0.001 |

<sup>a</sup>T<sub>0</sub> indicates the date of hypertension diagnosis; <sup>b</sup>T<sub>1</sub> indicates the date of first stroke admission or the matched outpatient visit for controls.
<sup>c</sup> Exposure variables are presented as median (interquartile range [IQR]) and compared using the Wilcoxon rank-sum test. Other continuous variables are presented as mean (SD) and compared using Welch's two-sample t-test; categorical variables are expressed as n (%) and compared using the Pearson chi-square test. Abbreviations: AF, atrial fibrillation; BP, blood pressure; COPD, chronic obstructive pulmonary disease; CHF, congestive heart failure; CKD, chronic kidney disease; HTN, hypertension; IQR, interquartile range; OP, outpatient visits; SVI, social vulnerability index.

During the matched observation window, cases had fewer outpatient visits with BP measurement per year than controls (median, 2.50 vs. 3.01; p < 0.001). Total number of antihypertensive medication ingredients had the same median in the two groups (median [IQR], 2.00 [1.00-4.00] in both groups). The intensification score defined using the 140/90 threshold was lower among cases than controls (median [IQR], -0.211 [-0.455, 0] vs. -0.125 [-0.333, 0]; p < 0.001), indicating a higher burden of elevated BP relative to medication intensification. Similar patterns were observed when the 160/100 threshold was used (median [IQR], 0 [0, 0.139] vs. 0 [0, 0.167]; p < 0.001). Distributions of the three hypertension care delivery measures are shown in Supplemental Figure 2.

### Association Between Hypertension Care Delivery and Ischemic Stroke Risk

In conditional logistic regression analyses, greater engagement in hypertension care delivery was generally associated with lower odds of ischemic stroke (Table 2).

**Table 2.**
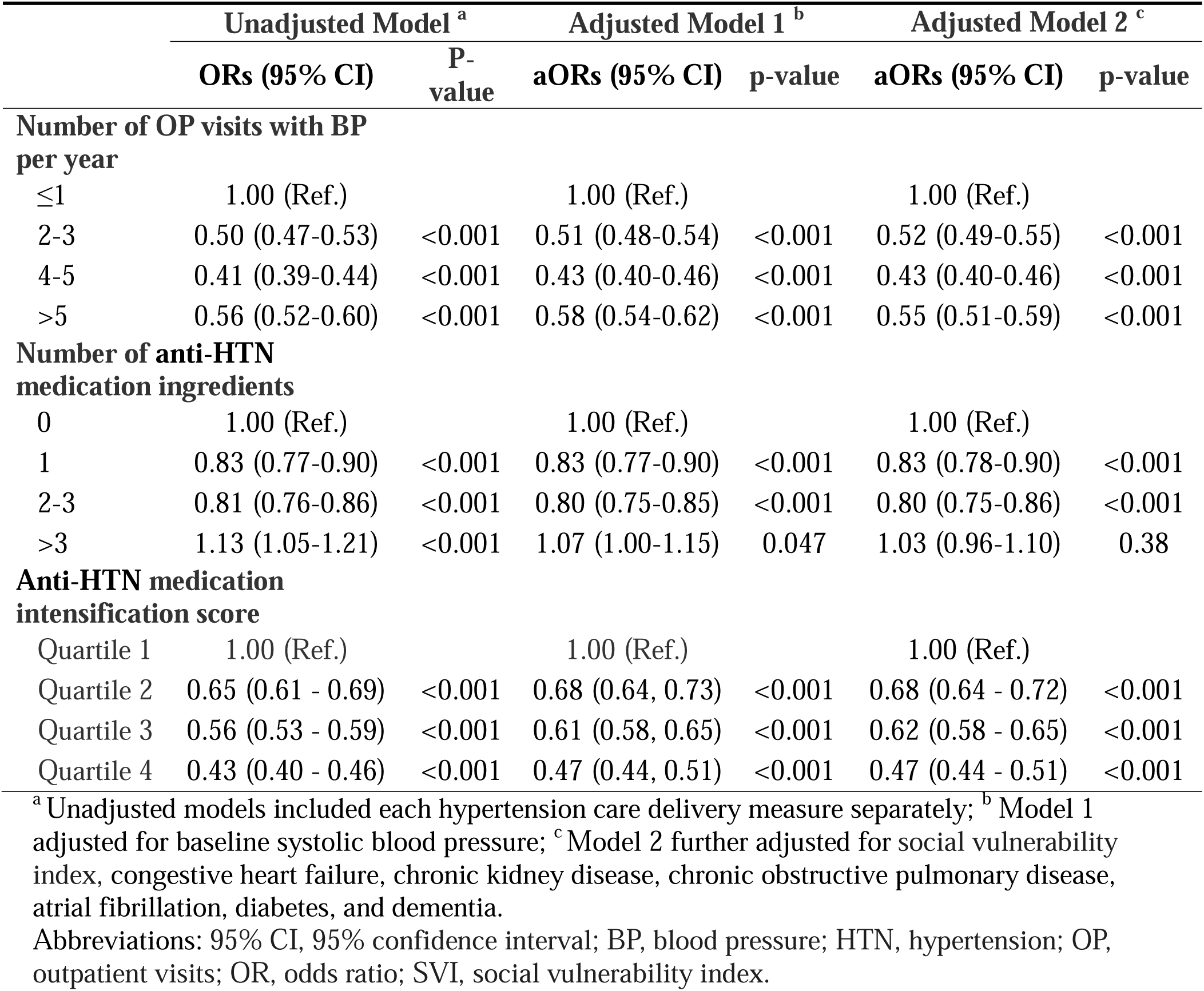
Conditional logistic regression of hypertension care delivery measures and stroke incidence.

|  | Unadjusted Model <sup>a</sup> |  | Adjusted Model 1 <sup>b</sup> |  | Adjusted Model 2 <sup>c</sup> |  |
| --- | --- | --- | --- | --- | --- | --- |
|  | ORs (95% CI) | P-value | aORs (95% CI) | p-value | aORs (95% CI) | p-value |
| **Number of OP visits with BP per year** |  |  |  |  |  |  |
| ≤1 | 1.00 (Ref.) |  | 1.00 (Ref.) |  | 1.00 (Ref.) |  |
| 2-3 | 0.50 (0.47-0.53) | <0.001 | 0.51 (0.48-0.54) | <0.001 | 0.52 (0.49-0.55) | <0.001 |
| 4-5 | 0.41 (0.39-0.44) | <0.001 | 0.43 (0.40-0.46) | <0.001 | 0.43 (0.40-0.46) | <0.001 |
| >5 | 0.56 (0.52-0.60) | <0.001 | 0.58 (0.54-0.62) | <0.001 | 0.55 (0.51-0.59) | <0.001 |
| **Number of anti-HTN medication ingredients** |  |  |  |  |  |  |
| 0 | 1.00 (Ref.) |  | 1.00 (Ref.) |  | 1.00 (Ref.) |  |
| 1 | 0.83 (0.77-0.90) | <0.001 | 0.83 (0.77-0.90) | <0.001 | 0.83 (0.78-0.90) | <0.001 |
| 2-3 | 0.81 (0.76-0.86) | <0.001 | 0.80 (0.75-0.85) | <0.001 | 0.80 (0.75-0.86) | <0.001 |
| >3 | 1.13 (1.05-1.21) | <0.001 | 1.07 (1.00-1.15) | 0.047 | 1.03 (0.96-1.10) | 0.38 |
| **Anti-HTN medication intensification score** |  |  |  |  |  |  |
| Quartile 1 | 1.00 (Ref.) |  | 1.00 (Ref.) |  | 1.00 (Ref.) |  |
| Quartile 2 | 0.65 (0.61 - 0.69) | <0.001 | 0.68 (0.64, 0.73) | <0.001 | 0.68 (0.64 - 0.72) | <0.001 |
| Quartile 3 | 0.56 (0.53 - 0.59) | <0.001 | 0.61 (0.58, 0.65) | <0.001 | 0.62 (0.58 - 0.65) | <0.001 |
| Quartile 4 | 0.43 (0.40 - 0.46) | <0.001 | 0.47 (0.44, 0.51) | <0.001 | 0.47 (0.44 - 0.51) | <0.001 |

<sup>a</sup> Unadjusted models included each hypertension care delivery measure separately; <sup>b</sup> Model 1 adjusted for baseline systolic blood pressure; <sup>c</sup> Model 2 further adjusted for social vulnerability index, congestive heart failure, chronic kidney disease, chronic obstructive pulmonary disease, atrial fibrillation, diabetes, and dementia.
Abbreviations: 95% CI, 95% confidence interval; BP, blood pressure; HTN, hypertension; OP, outpatient visits; OR, odds ratio; SVI, social vulnerability index.

For outpatient visits with BP measurement, increasing visit frequency up to 4-5 visits per year was associated with progressively lower odds of ischemic stroke. Compared with ≤1 visit per year, the adjusted odds ratios (aORs) in Model 2 were 0.52 (95% CI, 0.49-0.55) for 2-3 visits and 0.43 (95% CI, 0.40-0.46) for 4-5 visits. However, the association was attenuated among individuals with more than 5 visits per year (aOR, 0.55; 95% CI, 0.51-0.59), which may reflect greater underlying disease severity or higher healthcare utilization among patients requiring more frequent visits.

A similar pattern was observed for the number of antihypertensive medication ingredients. Compared with no antihypertensive medication use, the odds of stroke were lower among patients receiving one medication ingredient (aOR, 0.83; 95% CI, 0.78-0.90) and those receiving 2-3 ingredients (aOR, 0.80; 95% CI, 0.75-0.86). In contrast, use of more than three medication ingredients was not associated with lower stroke risk in fully adjusted models (aOR, 1.03; 95% CI, 0.96-1.10).

Higher treatment intensification scores were consistently associated with lower odds of ischemic stroke in a graded fashion. Compared with quartile 1, aORs in Model 2 were 0.68 (95% CI, 0.64-0.72) for quartile 2, 0.62 (95% CI, 0.58-0.65) for quartile 3, and 0.47 (95% CI, 0.44-0.51) for quartile 4. Results were similar in sensitivity analyses using a more stringent definition of elevated BP (≥160/100 mm Hg) (Supplemental Table 3).

## Subgroup Analyses by Baseline and Average SBP

Associations between hypertension care metrics and ischemic stroke were broadly consistent across strata of both baseline SBP and average SBP during the observation window (Figures 2 and 3). For outpatient visit frequency, more frequent visits were associated with lower odds of ischemic stroke across SBP strata, with no significant interaction by baseline SBP (p for interaction = 0.06) or average SBP (p for interaction = 0.38).

**Figure 2.**
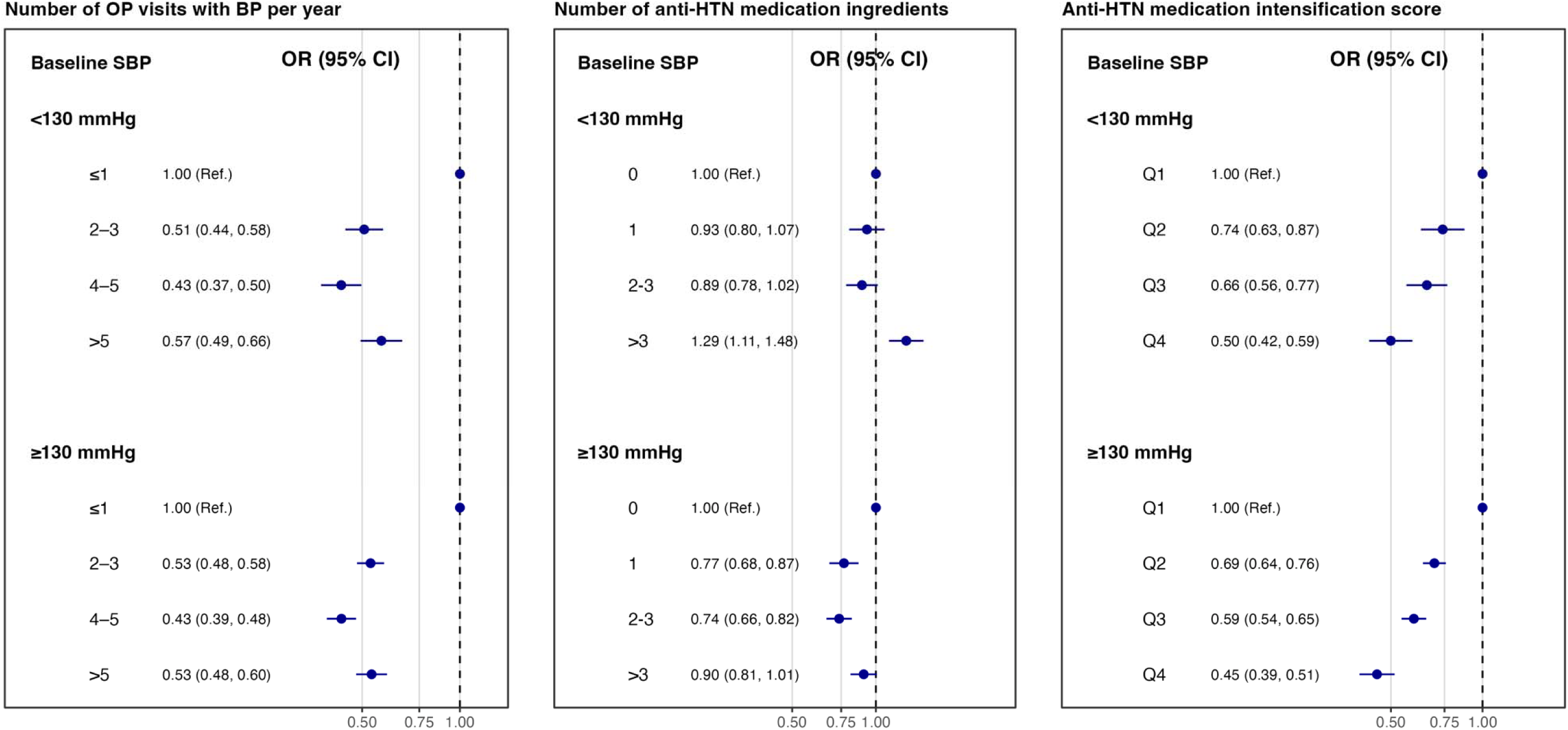
Association between hypertension care delivery measures and incident ischemic stroke stratified by baseline systolic blood pressure.

**Figure 3.**
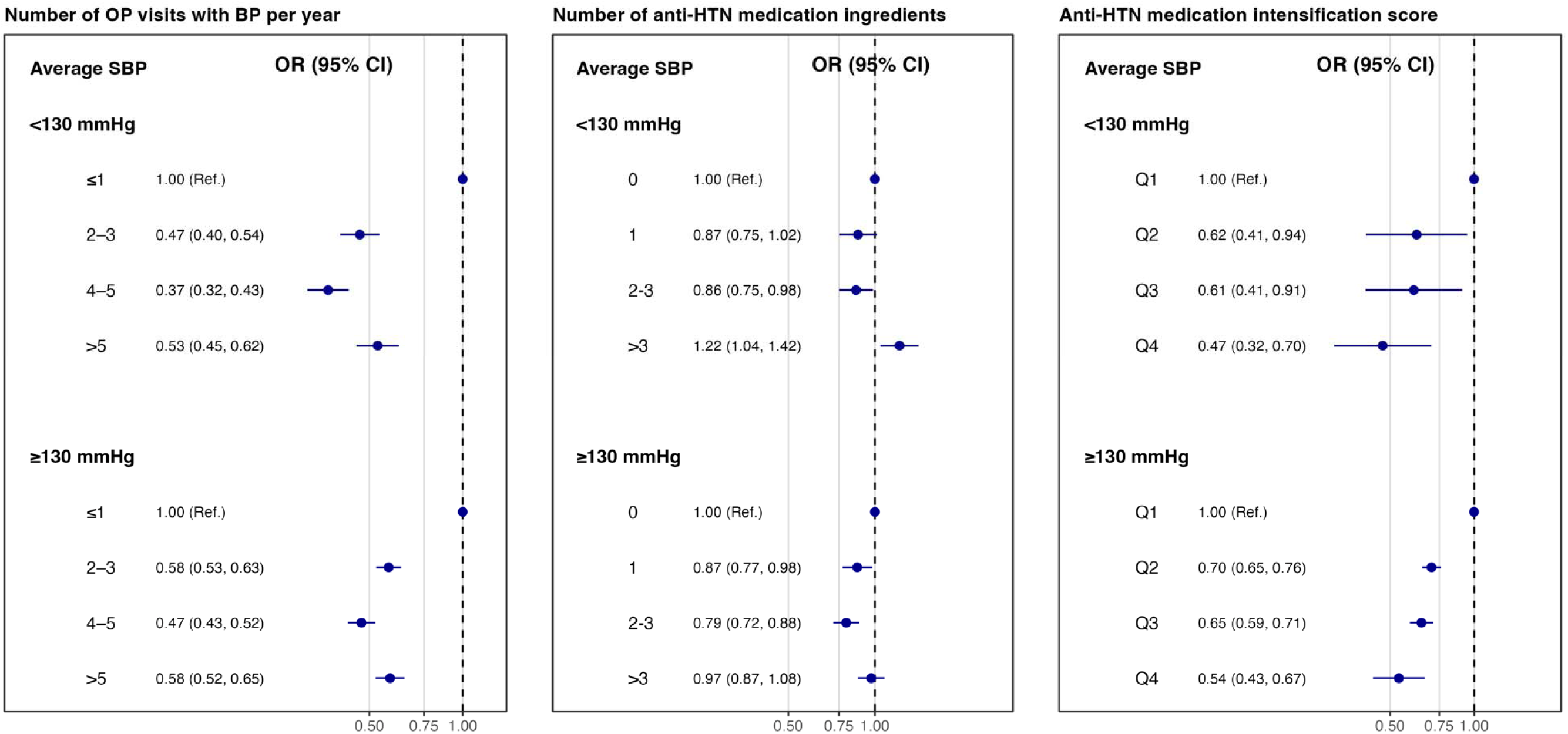
Association between hypertension care delivery measures and incident ischemic stroke stratified by baseline systolic blood pressure.

For the number of antihypertensive medication ingredients, the overall pattern of association with ischemic stroke was generally similar across strata of both baseline and average SBP. Although the interaction with baseline SBP reached statistical significance (p for interaction = 0.025), the direction of association was broadly consistent across SBP strata. In contrast, no interaction was observed by average SBP (p for interaction = 0.96).

For treatment intensification score, higher scores were associated with lower odds of ischemic stroke across SBP strata. However, both baseline SBP (p for interaction = 0.008) and average SBP (p for interaction = 0.041) showed modest effect modification, with somewhat stronger associations observed among individuals with higher SBP.

### Subgroup Analyses by Sex, Age, Race/Ethnicity, and SVI

Associations were similar between women and men (Supplemental Table 4). Compared with ≤1 outpatient BP visit per year, having 4-5 visits per year was associated with lower odds of ischemic stroke in both women (aOR 0.47, 95% CI 0.43-0.52) and men (aOR 0.38, 95% CI 0.35-0.42), with no evidence of interaction by sex (p for interaction = 0.13). Similarly, the associations for the number of antihypertensive medication ingredients and treatment intensification score did not differ significantly between sexes (p for interaction = 0.27 and 0.73, respectively).

In contrast, associations differed across age groups (Supplemental Table 5). The relationship between the number of antihypertensive medication ingredients and ischemic stroke risk varied significantly by age (p for interaction <0.001). In particular, use of >3 medication ingredients was associated with higher odds of ischemic stroke among individuals aged <45 years (aOR 1.74, 95% CI 1.29-2.36), whereas no clear association was observed among those aged ≥60 years (aOR 0.96, 95% CI 0.88-1.04). Effect modification by age was also observed for treatment intensification score (p for interaction <0.001).

Associations also varied by race and ethnicity (Supplemental Tables 6-1 and 6-2). Among non-Hispanic White and non-Hispanic Black patients, greater outpatient BP visit frequency and higher treatment intensification scores were associated with lower odds of ischemic stroke. However, estimates among Hispanic/Latino, non-Hispanic Asian, and other racial groups were less precise, likely reflecting smaller sample sizes. Significant interaction by race/ethnicity was observed for outpatient BP visit frequency and treatment intensification score (both p for interaction <0.001), whereas the association for medication ingredients counts did not differ significantly across racial and ethnic groups (p for interaction = 0.18).

Associations also varied across levels of social vulnerability (Supplemental Table 7). The association between outpatient BP visit frequency and ischemic stroke differed by SVI quartile (p for interaction = 0.011), with lower stroke odds associated with more frequent BP visits in lower SVI quartiles but a weaker association in the highest SVI quartile. Similarly, the relationship between the number of antihypertensive medication ingredients and ischemic stroke risk varied across SVI strata (p for interaction = 0.013). In contrast, the association between antihypertensive treatment intensification score and ischemic stroke risk was generally consistent across SVI quartiles (p for interaction = 0.18).

### Sensitivity Analyses

Sensitivity analyses excluding matched sets with baseline cardiometabolic conditions (CHF, AF, or CKD) yielded results similar to the primary analysis (Supplemental Table 8). In a more restrictive analysis excluding matched sets in which any patient had major baseline comorbidities (CHF, AF, CKD, COPD, diabetes, or dementia), the associations between outpatient BP visit frequency, antihypertensive medication ingredients, treatment intensification score, and ischemic stroke remained largely unchanged.

## Discussion

In this large, real-world, matched, case-control study of adults with hypertension, we identified patterns of outpatient hypertension care that were associated with incident ischemic stroke. Using complementary measures of BP monitoring frequency, antihypertensive medication use, and a treatment intensification score reflecting clinical responsiveness to elevated BP, we found that greater engagement in hypertension care was generally associated with lower odds of stroke. Specifically, more frequent outpatient BP visits and use of 1 to 3 antihypertensive medication ingredients were associated with lower stroke odds, and higher treatment intensification scores showed a graded inverse association with stroke risk. Importantly, these associations were observed across strata of both baseline SBP and average SBP during follow-up, suggesting that care delivery patterns capture aspects of hypertension management beyond BP level alone. These findings indicate that missed opportunities for ischemic stroke prevention may exist within routine hypertension care delivery.

Patterns of routine outpatient hypertension care appeared to play an important role in stroke risk. Patients with more frequent BP-related visits, particularly those with up to four to five visits per year, experienced substantially lower odds of ischemic stroke compared with individuals with minimal monitoring. Regular BP measurement may facilitate earlier detection of uncontrolled hypertension, timely treatment adjustments, and reinforcement of adherence to therapy and lifestyle modification.^13^ At the same time, the association between visit frequency and stroke risk did not show a consistent stepwise pattern across categories. Individuals with very frequent visits (>5 per year) did not demonstrate further reductions in stroke risk relative to those with four to five visits annually. This pattern likely reflects the complex relationship between care utilization and underlying disease severity. Patients requiring very frequent visits may represent a subgroup with more unstable BP, treatment-resistant hypertension, or greater comorbidity burden, factors that may partially offset the benefits of closer monitoring.^14,15^

The number of antihypertensive medication ingredients was also associated with stroke risk. Compared with patients receiving no antihypertensive medications, those treated with one or two to three medication ingredients had significantly lower odds of ischemic stroke, consistent with the established benefit of pharmacologic BP control in reducing cardiovascular risk.^16^ However, the use of more than three medication ingredients was not associated with additional reductions in stroke risk in fully adjusted models. This finding should be interpreted cautiously, as the number of prescribed medications may reflect both treatment intensity and underlying disease complexity.^16^ Patients receiving multiple antihypertensive agents often have more severe or treatment-resistant hypertension and may also have higher baseline cardiovascular risk.^17^ In this context, medication ingredient counts may simultaneously represent both intensified treatment and greater disease burden.

In contrast to the more heterogeneous patterns observed for visit frequency and medication ingredients counts, the treatment intensification score demonstrated a clear graded association with stroke risk. Higher intensification scores reflecting greater clinical responsiveness to elevated BP were consistently associated with lower odds of ischemic stroke. These findings are consistent with the concept of therapeutic inertia, defined as the failure to initiate or intensify antihypertensive therapy in the setting of uncontrolled BP.^18^ Prior studies have documented the widespread prevalence of therapeutic inertia across clinical settings and have linked it to poorer intermediate outcomes, particularly inadequate BP control.^6,19–21^ Our results extend this literature by demonstrating that patterns of treatment intensification over time may also be associated with differences in hard cardiovascular outcomes, including incident ischemic stroke.

Our findings are consistent with prior studies highlighting the importance of care processes in hypertension management. For example, Fontil et al. demonstrated that differences in treatment intensification and visit adherence were strongly associated with blood pressure control.^11^ In addition, large cohort and registry studies have shown that delays in medication intensification and prolonged intervals to follow-up after elevated BP are associated with increased risks of cardiovascular events or death, and lower treatment intensification scores are linked to higher risks of recurrent stroke.^22^ Building on this prior work, our study sought to characterize longitudinal patterns of hypertension care delivery across several complementary dimensions. In doing so, we demonstrate that care delivery patterns capture aspects of hypertension management beyond BP level alone.

Subgroup analyses provided additional insight into how these care delivery patterns may operate across patient populations. The association between outpatient BP monitoring frequency and stroke risk was broadly similar across both baseline SBP and average SBP during the observation window, suggesting that regular monitoring may confer benefits regardless of whether baseline BP is moderately elevated or more substantially elevated. In contrast, the relationships between medication ingredients counts and treatment intensification with stroke risk varied across certain demographic subgroups, including age and race or ethnicity. These differences may reflect variations in underlying hypertension severity, treatment response, comorbidity patterns, or healthcare access across populations. Importantly, however, the general direction of association for treatment intensification remained consistent across subgroups, reinforcing the importance of active clinical responsiveness to uncontrolled BP.

Taken together, these findings underscore the potential role of routine outpatient hypertension care processes in shaping long-term cardiovascular outcomes. Hypertension management in clinical practice often occurs through a series of outpatient encounters in which BP is measured, medications are adjusted, and adherence and lifestyle behaviors are discussed. Missed opportunities within these encounters, whether through infrequent monitoring, insufficient medication escalation, or delayed treatment intensification, may accumulate over time and contribute to the development of preventable cardiovascular events.^14,23,24^ From a pathophysiological perspective, chronic exposure to elevated BP contributes to endothelial dysfunction, arterial stiffening, and small-vessel disease, all of which increase susceptibility to ischemic stroke.^25–28^ These results suggest that improving the quality and responsiveness of routine hypertension care delivery may represent an important avenue for stroke prevention. Several practical strategies may help improve hypertension management in routine clinical practice, including structured follow-up programs to ensure regular BP monitoring, standardized protocols for BP measurement and treatment escalation, and the use of EHR-based clinical decision support tools to prompt timely medication adjustments. In addition, team-based care models involving pharmacists, nurses, and care coordinators may support medication management, patient education, and adherence, thereby improving long-term BP control.

This study has several strengths. First, it leveraged a large longitudinal EHR cohort within an integrated health system, enabling comprehensive capture of outpatient visits, BP measurements, and medication prescribing over extended follow-up. The integrated structure of the Sentara Health system, which includes hospitals, outpatient practices, and affiliated medical groups within a shared EHR environment, facilitates more complete longitudinal capture of routine hypertension care processes. Second, the matched case-control design enabled careful alignment of observation windows between cases and controls, reducing potential bias arising from differences in follow-up time. Third, we evaluated multiple complementary measures of hypertension care delivery, capturing different dimensions of outpatient management including monitoring, medication use, and treatment responsiveness. Together, these measures provide a more comprehensive representation of longitudinal hypertension care patterns than single cross-sectional metrics.

Several limitations should also be considered. First, this study was conducted within a single integrated regional health system. Although the large sample size and long observation period enhance the robustness of the findings, healthcare delivery models, insurance structures, and patient populations may differ across health systems and geographic regions. Although this regional health system is typical, the generalizability of these findings to other healthcare environments should be interpreted carefully. Second, as with all observational studies, residual confounding cannot be completely excluded, and the associations observed do not establish causality. Third, medication prescribing patterns were assessed using prescription records rather than measures of medication adherence, and actual medication use may therefore differ from recorded prescriptions. Fourth, care delivered outside the participating healthcare system may not have been captured, potentially leading to incomplete measurement of outpatient visits, medication exposure, or outcomes. Finally, the number of medication ingredients prescribed may reflect both treatment intensity and underlying disease severity, making interpretation of these associations complex.

In conclusion, longitudinal patterns of outpatient hypertension care delivery, including BP monitoring frequency, antihypertensive medication use, and treatment intensification, were associated with the risk of incident ischemic stroke in this large real-world cohort. These findings suggest that opportunities for stroke prevention may exist within routine outpatient hypertension care processes. Efforts to improve the consistency and responsiveness of hypertension management in everyday clinical practice—such as structured follow-up programs, standardized BP monitoring protocols, and timely treatment intensification supported by EHR-based decision support—may represent an important strategy for reducing the burden of stroke.

## Data Availability

Due to patient privacy considerations, the data underlying this study are housed within a secure environment maintained by Sentara Health and are not publicly available. However, limited deidentified data and additional details regarding the analytic methods may be made available by the corresponding author (J.B.) upon reasonable request.

## Acknowledgment

We gratefully acknowledge Dr. Valy Fontil for his expert guidance on the methodology for calculating the medication treatment intensification score. We also thank Dr. Jacob McPadden for his helpful input during the early stages of this study.

## Author Contributions

Y. Liu and J. Brush had full access to all of the study data and took full responsibility for the integrity and accuracy of the analysis. H. Yang, H. Krumholz, J. Brush, and Y. Lu are responsible for the conceptualization of the study. C. Kim, C. Huang, and M. Sawano contributed to the study design. P. Young, and M. Anderson contributed to the study analysis. H. Yang wrote the first draft, with contributions from all other authors. All authors reviewed and approved the final article.

## Nonstandard abbreviations and acronyms

AF: Atrial fibrillation
CHF: Congestive heart failure
CKD: Chronic kidney disease
COPD: Chronic obstructive pulmonary disease
DBP: Diastolic blood pressure
HER: Electronic health record
HTN: Hypertension
OMOP: Observational Medical Outcomes Partnership
OP: Outpatient
aOR: Adjusted odds ratio
RYV: Rate of years with visits
SBP: Systolic blood pressure
SVI: Social vulnerability index

