## Supplemental Material for "Missed Opportunities for Stroke Prevention in Hypertensive Patients: A Retrospective Case-Control Study"

**Supplemental Table 1. Definitions of clinical outcomes and comorbidities**.

| **Outcomes** | **Ancestor Concept IDs** |
| --- | --- |
| Hypertension | 320128, 4110948, 319826, 317895, 40481896, 43020424, 443771, 4311246, 45768449, 4316372, 4071202, 317898 |
| Myocardial infarction | 4270024, 4296653, 37309626, 312327, 43020460, 46270163, 4329847, 46270162, 45766241, 4108218, 45766114, 4108677, 438170, 444406, 438438, 441579, 434376, 436706 |
| Heart failure | 319835, 316139, 444101, 439696, 44782719, 40481043, 44782733, 40480602, 443587, 443580, 40480603, 40481042, 44782718, 4242669, 312927, 4273632, 439846, 35615055, 37309625, 4233424, 4004279, 4195785, 4264636, 4233224, 314378 |
| Ischemic stroke | 443454, 4108356, 4110189, 4043731, 45772786, 4110192, 4110190, 45767658, 46270031, 46273649, 40479572, 4046360, 4111714, 4045737, 4045738 |
| Cerebral hemorrhage | 376713 |
| Congestive heart failure | 319835 |
| Chronic kidney disease | 46271022 |
| Chronic obstructive pulmonary disease | 255573 |
| Atrial fibrillation | 313217 |
| Diabetes | 201820 |
| Dementia | 4182210 |

**Supplemental Table 2. Antihypertensive medications included in the study.**

| **Drug Class** | **Ingredient** | **Ancestor Concept ID** | **Drug Class** | **Ingredient** | **Ancestor Concept ID** |
| --- | --- | --- | --- | --- | --- |
| Thiazide-type diuretics | Chlorthalidone | 1395058 | Loop diuretics | Bumetanide | 932745 |
| Thiazide-type diuretics | Hydrochlorothiazide | 974166 | Loop diuretics | Furosemide | 956874 |
| Thiazide-type diuretics | Indapamide | 978555 | Loop diuretics | Torsemide | 942350 |
| ACE inhibitors | Benazepril | 1335471 | Potassium sparing diuretics | Amiloride | 991382 |
| ACE inhibitors | Captopril | 1340128 | Potassium sparing diuretics | Triamterene | 904542 |
| ACE inhibitors | Enalapril | 1341927 | Aldosterone antagonists | Eplerenone | 1309799 |
| ACE inhibitors | Fosinopril | 1363749 | Aldosterone antagonists | Spironolactone | 970250 |
| ACE inhibitors | Lisinopril | 1308216 | Beta blockers cardioselective | Atenolol | 1314002 |
| ACE inhibitors | Moexipril | 1310756 | Beta blockers cardioselective | Betaxolol | 1322081 |
| ACE inhibitors | Perindopril | 1373225 | Beta blockers cardioselective | Bisoprolol | 1338005 |
| ACE inhibitors | Quinapril | 1331235 | Beta blockers cardioselective | Metoprolol | 1307046 |
| ACE inhibitors | Ramipril | 1334456 | Beta blockers vasodilatory | Nebivolol | 1314577 |
| ACE inhibitors | Trandolapril | 1342439 | Beta blockers noncardioselective | Nadolol | 1313200 |
| ARBs | Azilsartan | 40235485 | Beta blockers noncardioselective | Propranolol | 1353766 |
| ARBs | Candesartan | 1351557 | Beta blockers ISA | Acebutolol | 1319998 |
| ARBs | Eprosartan | 1346686 | Beta blockers ISA | Penbutolol | 1327978 |
| ARBs | Irbesartan | 1347384 | Beta blockers ISA | Pindolol | 1345858 |
| ARBs | Losartan | 1367500 | Alpha Beta blockers | Carvedilol | 1346823 |
| ARBs | Olmesartan | 40226742 | Alpha Beta blockers | Labetalol | 1386957 |
| ARBs | Telmisartan | 1317640 | Direct renin inhibitor | Aliskiren | 1317967 |
| ARBs | Valsartan | 1308842 | Alpha1 blockers | Doxazosin | 1363053 |
| CCB dihydropyridines | Amlodipine | 1332418 | Alpha1 blockers | Prazosin | 1350489 |
| CCB dihydropyridines | Felodipine | 1353776 | Alpha1 blockers | Terazosin | 1341238 |
| CCB dihydropyridines | Isradipine | 1326012 | Central alpha2 agonists | Clonidine | 1398937 |
| CCB dihydropyridines | Nicardipine | 1318137 | Central alpha2 agonists | Methyldopa | 1305447 |
| CCB dihydropyridines | Nifedipine | 1318853 | Central alpha2 agonists | Guanfacine | 1344965 |
| CCB dihydropyridines | Nisoldipine | 1319880 | Direct vasodilators | Hydralazine | 1373928 |
| CCB nondihydropyridines | Diltiazem | 1328165 | Direct vasodilators | Minoxidil | 1309068 |
| CCB nondihydropyridines | Verapamil | 1307863 | Dual endothelin receptor antagonist | Aprocitentan | 42609551 |

**Supplemental Table 3. Sensitivity analysis of antihypertensive medication intensification score defined using a 160/100 mmHg threshold.**

|  | **Unadjusted Model** | | **Adjusted Model 1^a^** | | **Adjusted Model 2^b^** | |
| --- | --- | --- | --- | --- | --- | --- |
|  | **ORs (95% CI)** | **p-value** | **aORs (95% CI)** | **p-value** | **aORs (95% CI)** | **p-value** |
| Quartile 1 | 1.00 (Ref.) |  | 1.00 (Ref.) |  | 1.00 (Ref.) |  |
| Quartile 2 | 0.64 (0.58-0.70) | <0.001 | 0.68 (0.62-0.74) | <0.001 | 0.65 (0.59-0.72) | <0.001 |
| Quartile 3 | 0.59 (0.55-0.62) | <0.001 | 0.62 (0.59-0.66) | <0.001 | 0.62 (0.58-0.65) | <0.001 |
| Quartile 4 | 0.57 (0.54-0.61) | <0.001 | 0.62 (0.58-0.65) | <0.001 | 0.62 (0.59-0.66) | <0.001 |

^a^ Model 1 adjusted for baseline systolic blood pressure; ^b^ Model 2 further adjusted for social vulnerability index, congestive heart failure, chronic kidney disease, chronic obstructive pulmonary disease, atrial fibrillation, diabetes, and dementia.

Abbreviations: 95% CI, 95% confidence interval; OR, odds ratio.

**Supplemental Table 4. Subgroup analyses stratified by sex**.

|  | **Adjusted Model** | | | | **p-interaction** |
| --- | --- | --- | --- | --- | --- |
|  | **Females** | | **Males** |  |  |
|  | **aORs (95% CI)** | **p-value** | **aORs (95% CI)** | **p-value** |  |
| **Number of OP visits with BP per year** |  |  |  |  | 0.13 |
| ≤1 | 1.00 (Ref.) |  | 1.00 (Ref.) |  |  |
| 2-3 | 0.56 (0.52-0.61) | <0.001 | 0.48 (0.43-0.52) | <0.001 |  |
| 4-5 | 0.47 (0.43-0.52) | <0.001 | 0.38 (0.35-0.42) | <0.001 |  |
| >5 | 0.57 (0.52-0.63) | <0.001 | 0.52 (0.47-0.58) | <0.001 |  |
| **Number of anti-HTN medications ingredients** |  |  |  |  | 0.27 |
| 0 | 1.00 (Ref.) |  | 1.00 (Ref.) |  |  |
| 1 | 0.88 (0.80-0.98) | 0.016 | 0.79 (0.71-0.88) | <0.001 |  |
| 2-3 | 0.82 (0.75-0.90) | <0.001 | 0.78 (0.71-0.86) | <0.001 |  |
| >3 | 1.07 (0.98-1.18) | 0.14 | 0.98 (0.89-1.09) | 0.77 |  |
| **Anti-HTN medication intensification ratio** |  |  |  |  | 0.73 |
| Quartile 1 | 1.00 (Ref.) |  | 1.00 (Ref.) |  |  |
| Quartile 2 | 0.70 (0.64, 0.76) | <0.001 | 0.66 (0.61, 0.73) | <0.001 |  |
| Quartile 3 | 0.62 (0.57, 0.67) | <0.001 | 0.61 (0.56, 0.67) | <0.001 |  |
| Quartile 4 | 0.48 (0.44, 0.53) | <0.001 | 0.46 (0.41, 0.51) | <0.001 |  |

Model adjusted for baseline systolic blood pressure, congestive heart failure, chronic kidney disease, chronic obstructive pulmonary disease, atrial fibrillation, diabetes, and dementia.

Abbreviations: 95% CI, 95% confidence interval; BP, blood pressure; HTN, hypertension; OP, outpatient visits; OR, odds ratio.

**Supplemental Table 5. Subgroup analyses stratified by age group.**

|  | **Adjusted Model** | | | | | | **p-interaction** |
| --- | --- | --- | --- | --- | --- | --- | --- |
|  | **< 45 years** | | **45-59 years** | | **≥ 60 years** | |  |
|  | **aORs (95% CI)** | **p-value** | **aORs (95% CI)** | **p-value** | **aORs (95% CI)** | **p-value** |  |
| **Number of OP visits with BP per year** |  |  |  |  |  |  | 0.75 |
| ≤1 | 1.00 (Ref.) |  | 1.00 (Ref.) |  | 1.00 (Ref.) |  |  |
| 2-3 | 0.43 (0.34-0.55) | <0.001 | 0.50 (0.45-0.56) | <0.001 | 0.54 (0.5-0.59) | <0.001 |  |
| 4-5 | 0.39 (0.3-0.52) | <0.001 | 0.41 (0.36-0.47) | <0.001 | 0.44 (0.4-0.48) | <0.001 |  |
| >5 | 0.6 (0.43-0.83) | 0.002 | 0.60 (0.52-0.68) | <0.001 | 0.53 (0.49-0.58) | <0.001 |  |
| **Number of anti-HTN medications ingredients** |  |  |  |  |  |  | <0.001 |
| 0 | 1.00 (Ref.) |  | 1.00 (Ref.) |  | 1.00 (Ref.) |  |  |
| 1 | 0.85 (0.61-1.19) | 0.35 | 0.81 (0.70-0.94) | 0.004 | 0.84 (0.77-0.92) | <0.001 |  |
| 2-3 | 0.97 (0.73-1.29) | 0.84 | 0.81 (0.71-0.93) | 0.002 | 0.79 (0.73-0.85) | <0.001 |  |
| >3 | 1.74 (1.29-2.36) | 0.001 | 1.12 (0.97-1.28) | 0.11 | 0.96 (0.88-1.04) | 0.28 |  |
| **Anti-HTN medication intensification ratio** |  |  |  |  |  |  | <0.001 |
| Quartile 1 | 1.00 (Ref.) |  | 1.00 (Ref.) |  | 1.00 (Ref.) |  |  |
| Quartile 2 | 0.58 (0.44-0.75) | <0.001 | 0.62 (0.55-0.7) | <0.001 | 0.72 (0.67-0.77) | <0.001 |  |
| Quartile 3 | 0.46 (0.35-0.59) | <0.001 | 0.56 (0.5-0.63) | <0.001 | 0.66 (0.61-0.71) | <0.001 |  |
| Quartile 4 | 0.44 (0.32-0.61) | <0.001 | 0.39 (0.34-0.45) | <0.001 | 0.51 (0.47-0.56) | <0.001 |  |

Model adjusted for baseline systolic blood pressure, congestive heart failure, chronic kidney disease, chronic obstructive pulmonary disease, atrial fibrillation, diabetes, and dementia.

Abbreviations: 95% CI, 95% confidence interval; BP, blood pressure; HTN, hypertension; OP, outpatient visits; OR, odds ratio.

**Supplemental Table 6-1. Subgroup analyses stratified by race and ethnicity.**

|  | **Adjusted Model** | | | | | |
| --- | --- | --- | --- | --- | --- | --- |
|  | **Non-Hispanic White** | | **Non-Hispanic Black** | | **Non-Hispanic Asian** | |
|  | **aORs (95% CI)** | **p-value** | **aORs (95% CI)** | **p-value** | **aORs (95% CI)** | **p-value** |
| **Number of OP visits with BP per year** |  |  |  |  |  |  |
| ≤1 | 1.00 (Ref.) |  | 1.00 (Ref.) |  | 1.00 (Ref.) |  |
| 2-3 | 0.52 (0.48-0.56) | <0.001 | 0.52 (0.47-0.57) | <0.001 | 0.58 (0.23-1.44) | 0.24 |
| 4-5 | 0.44 (0.41-0.48) | <0.001 | 0.4 (0.36-0.45) | <0.001 | 0.71 (0.27-1.88) | 0.50 |
| >5 | 0.58 (0.53-0.63) | <0.001 | 0.48 (0.43-0.55) | <0.001 | 0.74 (0.26-2.11) | 0.58 |
| **Number of anti-HTN medications ingredients** |  |  |  |  |  |  |
| 0 | 1.00 (Ref.) |  | 1.00 (Ref.) |  | 1.00 (Ref.) |  |
| 1 | 0.84 (0.77-0.92) | <0.001 | 0.83 (0.72-0.95) | 0.008 | 0.67 (0.18-2.52) | 0.55 |
| 2-3 | 0.85 (0.78-0.92) | <0.001 | 0.72 (0.63-0.81) | <0.001 | 0.99 (0.33-2.98) | 0.98 |
| >3 | 1.11 (1.02-1.21) | 0.016 | 0.89 (0.79-1.01) | 0.062 | 1.64 (0.5-5.41) | 0.42 |
| **Anti-HTN medication intensification ratio** |  |  |  |  |  |  |
| Quartile 1 | 1.00 (Ref.) |  | 1.00 (Ref.) |  | 1.00 (Ref.) |  |
| Quartile 2 | 0.72 (0.67-0.78) | <0.001 | 0.63 (0.57-0.7) | <0.001 | 0.89 (0.37-2.1) | 0.78 |
| Quartile 3 | 0.66 (0.61-0.71) | <0.001 | 0.55 (0.49-0.61) | <0.001 | 0.86 (0.35-2.11) | 0.74 |
| Quartile 4 | 0.53 (0.48-0.58) | <0.001 | 0.37 (0.32-0.42) | <0.001 | 0.71 (0.26-1.92) | 0.50 |

Model adjusted for baseline systolic blood pressure, congestive heart failure, chronic kidney disease, chronic obstructive pulmonary disease, atrial fibrillation, diabetes, and dementia.

Abbreviations: 95% CI, 95% confidence interval; BP, blood pressure; HTN, hypertension; OP, outpatient visits; OR, odds ratio.

**Supplemental Table 6-2.** **Subgroup analyses stratified by race and ethnicity.**

|  | **Adjusted Model** | | | | **p-interaction** |
| --- | --- | --- | --- | --- | --- |
|  | **Hispanic/Latino** | | **Other/Unknown** | |  |
|  | **aORs (95% CI)** | **p-value** | **aORs (95% CI)** | **p-value** |  |
| **Number of OP visits with BP per year** |  |  |  |  | <0.001 |
| ≤1 | 1.00 (Ref.) |  | 1.00 (Ref.) |  |  |
| 2-3 | 0.63 (0.21-1.88) | 0.40 | 1.69 (0.42-6.78) | 0.46 |  |
| 4-5 | 0.39 (0.11-1.39) | 0.15 | 1.2 (0.25-5.69) | 0.82 |  |
| >5 | 0.32 (0.08-1.21) | 0.09 | 5.39 (0.89-32.65) | 0.07 |  |
| **Number of anti-HTN medications ingredients** |  |  |  |  | 0.18 |
| 0 | 1.00 (Ref.) |  | 1.00 (Ref.) |  |  |
| 1 | 1.33 (0.37-4.8) | 0.66 | 1.22 (0.16-9.16) | 0.84 |  |
| 2-3 | 0.76 (0.26-2.19) | 0.61 | 0.55 (0.08-3.82) | 0.54 |  |
| >3 | 0.85 (0.24-3.03) | 0.80 | 1.24 (0.18-8.47) | 0.83 |  |
| **Anti-HTN medication intensification ratio** |  |  |  |  | <0.001 |
| Quartile 1 | 1.00 (Ref.) |  | 1.00 (Ref.) |  |  |
| Quartile 2 | 1.3 (0.36-4.66) | 0.69 | 0.59 (0.16-2.11) | 0.42 |  |
| Quartile 3 | 0.7 (0.22-2.24) | 0.55 | 0.48 (0.12-1.89) | 0.29 |  |
| Quartile 4 | 1.00 (0.28-3.6) | >0.99 | 0.3 (0.05-1.76) | 0.18 |  |

Model adjusted for baseline systolic blood pressure, congestive heart failure, chronic kidney disease, chronic obstructive pulmonary disease, atrial fibrillation, diabetes, and dementia.

Abbreviations: 95% CI, 95% confidence interval; BP, blood pressure; HTN, hypertension; OP, outpatient visits; OR, odds ratio.

**Supplemental Table 7. Subgroup analyses stratified by social vulnerability index.**

|  | **Adjusted Model** | | | | | | | | **p-interaction** |
| --- | --- | --- | --- | --- | --- | --- | --- | --- | --- |
|  | **Quartile 1** | | **Quartile 2** | | **Quartile 3** | | **Quartile 4** | |  |
|  | **aORs (95% CI)** | **p-value** | **aORs (95% CI)** | **p-value** | **aORs (95% CI)** | **p-value** | **aORs (95% CI)** | **p-value** |  |
| **Number of OP visits with BP per year** |  |  |  |  |  |  |  |  | 0.011 |
| ≤1 | 1.00 (Ref.) |  | 1.00 (Ref.) |  | 1.00 (Ref.) |  | 1.00 (Ref.) |  |  |
| 2-3 | 0.49 (0.39-0.61) | <0.001 | 0.53 (0.42-0.66) | <0.001 | 0.56 (0.43-0.74) | <0.001 | 0.95 (0.66-1.36) | 0.79 |  |
| 4-5 | 0.41 (0.32-0.52) | <0.001 | 0.53 (0.42-0.68) | <0.001 | 0.36 (0.26-0.49) | <0.001 | 0.58 (0.39-0.85) | 0.005 |  |
| >5 | 0.61 (0.47-0.77) | <0.001 | 0.51 (0.39-0.66) | <0.001 | 0.54 (0.39-0.75) | <0.001 | 0.59 (0.39-0.89) | 0.012 |  |
| **Number of anti-HTN medications ingredients** |  |  |  |  |  |  |  |  | 0.013 |
| 0 | 1.00 (Ref.) |  | 1.00 (Ref.) |  | 1.00 (Ref.) |  | 1.00 (Ref.) |  |  |
| 1 | 1.11 (0.87-1.41) | 0.41 | 0.76 (0.58-0.99) | 0.045 | 0.63 (0.45-0.89) | 0.009 | 0.62 (0.38-1.02) | 0.058 |  |
| 2-3 | 1.02 (0.82-1.26) | 0.89 | 0.95 (0.74-1.21) | 0.66 | 0.79 (0.58-1.06) | 0.12 | 0.58 (0.38-0.88) | 0.011 |  |
| >3 | 1.37 (1.08-1.73) | 0.008 | 1.06 (0.82-1.37) | 0.67 | 0.87 (0.64-1.18) | 0.38 | 0.87 (0.57-1.33) | 0.52 |  |
| **Anti-HTN medication intensification ratio** |  |  |  |  |  |  |  |  | 0.18 |
| Quartile 1 | 1.00 (Ref.) |  | 1.00 (Ref.) |  | 1.00 (Ref.) |  | 1.00 (Ref.) |  |  |
| Quartile 2 | 0.73 (0.59-0.91) | 0.006 | 0.75 (0.60-0.93) | 0.011 | 0.84 (0.64-1.11) | 0.22 | 0.54 (0.37-0.77) | 0.001 |  |
| Quartile 3 | 0.62 (0.50-0.77) | <0.001 | 0.80 (0.64-0.99) | 0.044 | 0.64 (0.48-0.85) | 0.002 | 0.50 (0.34-0.73) | <0.001 |  |
| Quartile 4 | 0.49 (0.38-0.63) | <0.001 | 0.55 (0.41-0.72) | <0.001 | 0.58 (0.41-0.82) | 0.002 | 0.38 (0.24-0.63) | <0.001 |  |

Model adjusted for baseline systolic blood pressure, congestive heart failure, chronic kidney disease, chronic obstructive pulmonary disease, atrial fibrillation, diabetes, and dementia.

Abbreviations: 95% CI, 95% confidence interval; BP, blood pressure; HTN, hypertension; OP, outpatient visits; OR, odds ratio; SVI, social vulnerability index.

**Supplemental Table 8. Sensitivity analyses excluding matched sets with baseline cardiometabolic conditions or major comorbidities.**

|  | **Adjusted Model** | | | |
| --- | --- | --- | --- | --- |
|  | **Cardiometabolic exclusion ^a^** | | **Major comorbidity exclusion ^b^** | |
|  | **aORs (95% CI)** | **p-value** | **aORs (95% CI)** | **p-value** |
| **Number of OP visits with BP per year** |  |  |  |  |
| ≤1 | 1.00 (Ref.) |  | 1.00 (Ref.) |  |
| 2-3 | 0.51 (0.47-0.54) | <0.001 | 0.45 (0.40-0.50) | <0.001 |
| 4-5 | 0.41 (0.38-0.45) | <0.001 | 0.38 (0.34-0.43) | <0.001 |
| >5 | 0.55 (0.51-0.59) | <0.001 | 0.50 (0.44-0.57) | <0.001 |
| **Number of anti-HTN medications ingredients** |  |  |  |  |
| 0 | 1.00 (Ref.) |  | 1.00 (Ref.) |  |
| 1 | 0.83 (0.77-0.91) | <0.001 | 0.76 (0.66-0.87) | <0.001 |
| 2-3 | 0.80 (0.74-0.86) | <0.001 | 0.79 (0.71-0.89) | <0.001 |
| >3 | 1.05 (0.97-1.14) | 0.21 | 1.04 (0.91-1.18) | 0.59 |
| **Anti-HTN medication intensification ratio** |  |  |  |  |
| Quartile 1 | 1.00 (Ref.) |  | 1.00 (Ref.) |  |
| Quartile 2 | 0.68 (0.63-0.72) | <0.001 | 0.68 (0.60-0.76) | <0.001 |
| Quartile 3 | 0.60 (0.56-0.64) | <0.001 | 0.61 (0.54-0.68) | <0.001 |
| Quartile 4 | 0.45 (0.41-0.49) | <0.001 | 0.48 (0.42-0.55) | <0.001 |

^a^ Models were adjusted for baseline systolic blood pressure, social vulnerability index, chronic obstructive pulmonary disease, diabetes, and dementia.

^b^ Models were adjusted for baseline systolic blood pressure and social vulnerability index.

Abbreviations: 95% CI, 95% confidence interval; BP, blood pressure; HTN, hypertension; OP, outpatient visits; OR, odds ratio.

**Supplemental Figure 1. Flow chart of participants.**

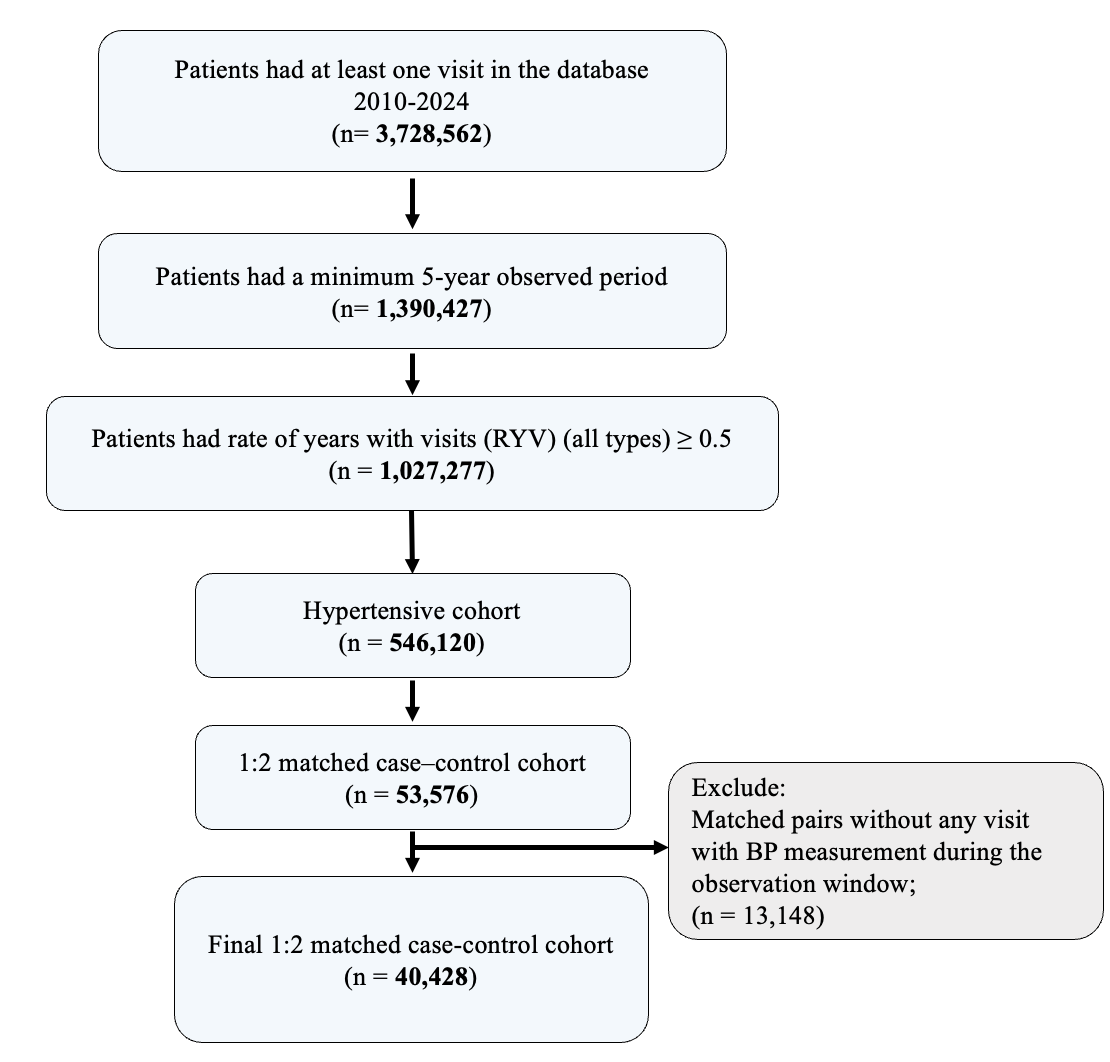

**Supplemental Figure 2. Distribution of hypertension care delivery measures among case and control groups.**

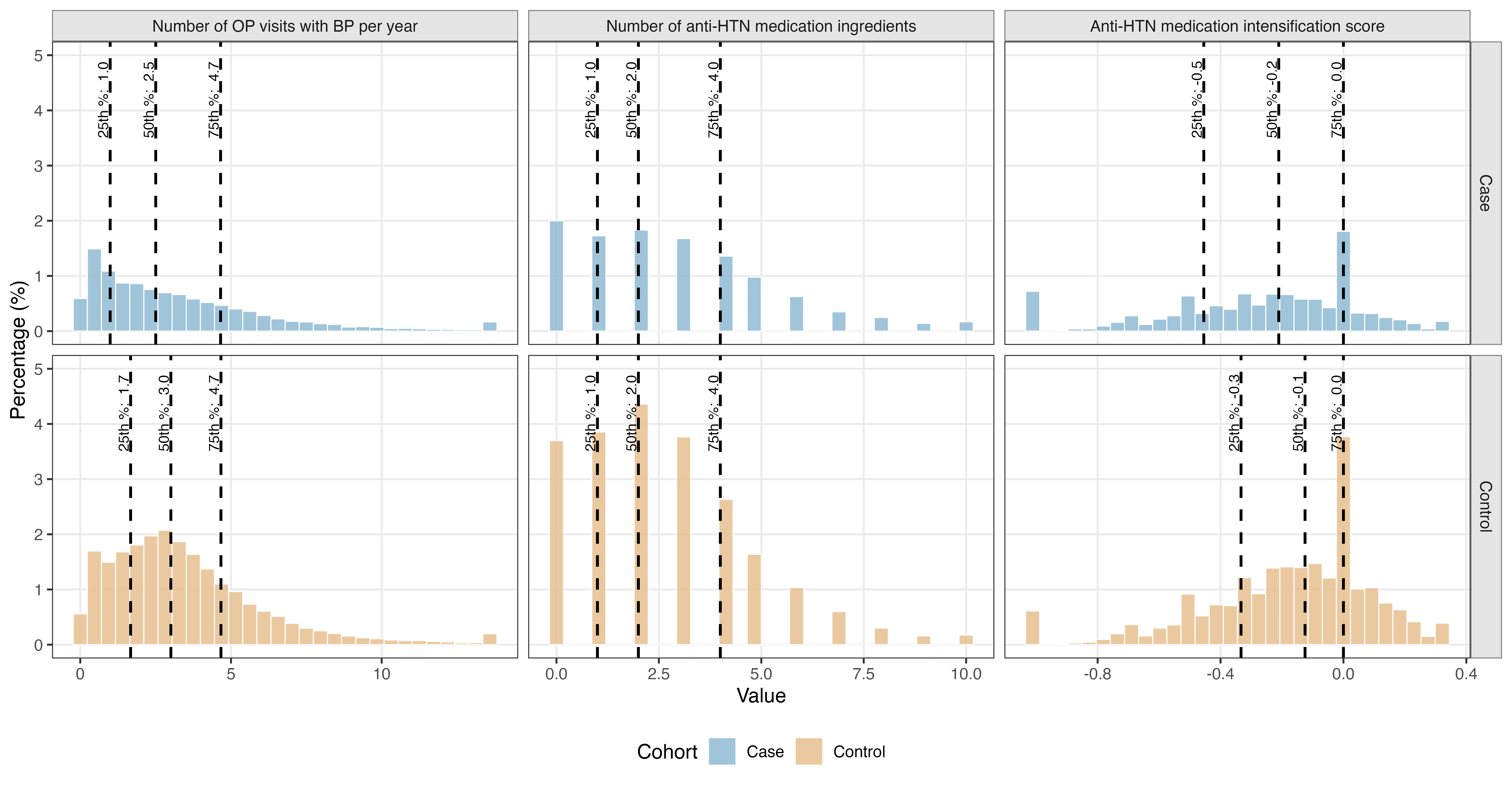
